# Joint modelling of PSA dynamics and prostate cancer risks: A population-based study

**DOI:** 10.64898/2026.02.15.26346131

**Authors:** Bizhan Akynkozhayev, Benjamin Christoffersen, Anna Lantz, Tobias Nordström, Keith Humphreys, Mark Clements

**Author notes:** Corresponding author: (BA). These authors contributed equally to this work (Bizhan Akynkozhayev, Benjamin Christoffersen, Keith Humphreys, Mark Clements). These authors also contributed equally to this work (Anna Lantz, Tobias Nordström).

## Abstract

While the prostate-specific antigen (PSA) test is a widely used prostate cancer screening tool, its application remains controversial. Opportunistic PSA testing generates complex data in which testing intensities, PSA levels, and prostate cancer diagnosis are interdependent. Conventional analyses rarely model these processes jointly. The objective of this study was to develop a population-based joint model to analyse PSA dynamics, retesting patterns, and prostate cancer risk. We used the Stockholm Prostate Cancer Diagnostics Register to identify 506,761 men with at least one PSA test between 2003 and 2020. We fitted a joint model linking three components: a linear mixed-effects submodel for PSA over age, and two proportional hazards submodels for time to next PSA test and time to prostate cancer diagnosis. PSA increased nonlinearly with age, with substantial between-person heterogeneity and increasing unexplained variation with increasing age. In the joint model, doubling of the total PSA values was associated with a hazard ratio (HR) of 2.01 (95% CI: 1.99–2.02; P < .001) for diagnosis and 1.163 (95% CI: 1.161–1.165; P < .001) for retesting. These hazard ratios were significantly stronger than estimates obtained when modelling these processes separately. As a limitation, the study is primarily limited by its observational nature and a lack of data on non-cancer factors that can elevate PSA, such as urinary tract infections or lower urinary tract symptoms. Furthermore, the model does not explicitly account for PSA trajectory changes after cancer onset. In conclusion, jointly modelling PSA dynamics and testing behaviour corrects for the informative observation bias inherent in opportunistic testing. This approach yields more accurate population estimates and personalised risk predictions compared to traditional isolated models. Our findings suggest that PSA dynamics may be clinically informative and that screening models should jointly incorporate testing history and PSA trajectories to improve precision.

## Introduction

Prostate cancer is the most common cancer and is the third leading cause of cancer-related death among men in the European Union [1]. PSA is an inexpensive blood test that has long guided decisions on prostate cancer screening. The use of a single PSA threshold remains controversial [2–4]. The European Randomized Study of Screening for Prostate Cancer [5] and the Cluster Randomized Trial of PSA Testing for Prostate Cancer [6] demonstrated significant reductions in prostate cancer mortality, whereas the U.S. Prostate, Lung, Colorectal and Ovarian trial found no such reduction [7]. PSA levels increase with age and age-specific thresholds have been proposed to improve screening accuracy, yet their use remains controversial [8–10].

Longitudinal PSA trajectories may provide information beyond a single measurement. PSA doubling times have been linked to aggressive disease [11,12], although population-level studies have questioned their predictive value [13,14].

Large trials and health-system cohorts have documented extensive repeat PSA testing in routine care [15–20], but most analyses have considered only one of three key processes: longitudinal PSA trajectories, testing patterns (the observational process), or time to prostate cancer diagnosis. Few studies have applied joint modelling frameworks, which link longitudinal measurements with time-to-event outcomes, to PSA data, but these have focused on men after a prostate cancer diagnosis [21–23]. There is limited evidence on how PSA trajectories, repeat testing, and diagnosis are inter-related in men preceding any diagnosis of prostate cancer. Moreover, existing studies have not explicitly accounted for the fact that men with higher PSA values are more likely to undergo frequent testing. Recent computational developments now make a joint analysis of all three processes feasible for larger register-based datasets.

In this study, we develop a detailed population-based PSA model. Using extensive PSA test data linked with rich health and population registers, we describe: (a) PSA trajectories, which are allowed to vary between individuals; (b) time to prostate cancer diagnosis; and (c) PSA testing patterns, with both diagnosis and testing dependent on the underlying PSA trajectory. Jointly modelling these longitudinal and time-to-event processes may provide novel insights into both the PSA dynamics and the rescreening behaviour. Specifically, we expect that predictions from the joint model will more accurately capture the underlying data-generating mechanisms than those from separate models. Such a model could be used to predict whether a man’s PSA levels are rising faster than expected of their age, signalling the need for closer clinical follow-up.

## Materials and methods

### Materials

This study uses data from the Stockholm Prostate Cancer Diagnostics Register [24], a comprehensive regional database that links information from Swedish national and regional registers, including PSA test dates and results, biopsies, cancer diagnoses, cause of death, and population data for men residing in Stockholm County. These data reflect a healthcare system where organised prostate cancer screening has not been recommended by the Swedish Board of Health and Welfare. Men may instead discuss prostate cancer testing with their clinician through a process of shared decision-making. For men who choose to have a PSA test, the Swedish national guidelines (Nationellt vårdprogram för prostatacancer) suggest that a raised PSA test is followed by magnetic resonance imaging (MRI) and then possibly a referral for a biopsy. The specific age-dependent PSA thresholds for referral into a standardised care pathway (Standardiserat vårdförlopp) are ≥ 3.0 ng/mL for men under 70 years, ≥ 5.0 ng/mL for those aged 70–80, and ≥ 7.0 ng/mL for those over 80. In clinical practice, if an initial PSA result is between 3 and 10 ng/mL and the prostate palpation is benign, a second PSA test is recommended after approximately three weeks to account for intra-individual variation before proceeding to MRI. Further, the indications for biopsy are refined using PSA density; for example, a biopsy is recommended for PI-RADS 3 findings only if the PSA density is ≥ 0.10 ng/mL/cm^3^, or for PI-RADS 1–2 findings if the density is ≥ 0.20 ng/mL/cm^3^.

The data were accessed on 12 January 2023 under the ethical approval from the Regional Ethical Review Board in Stockholm (Dnr 2012/438-31/3), with a subsequent amendment (Dnr 2016/620-32). As this was a register-based study, the requirement for individual informed consent was waived by the ethics committee. All data were processed anonymously, and the authors had no access to information that could be used to identify individual participants during or after the study.

The study population comprises 506,761 men who underwent at least one PSA test between 2003 and 2020, with a median follow-up of 9.8 years and a total of 1,836,900 tests recorded during this period. There was a median of two tests per man, and 303,752 men had at least two tests. Disease verification was register-based and relied on biopsy-confirmed diagnoses, with predominantly systematic biopsies before 2018 and a rapidly increasing use of pre-biopsy MRI thereafter. PSA values and testing patterns change drastically after diagnosis and treatment, and the post-diagnosis is not the focus of this study; we therefore censored follow-up at the date of prostate cancer diagnosis and excluded 335,172 tests taken after the diagnosis date. After censoring for diagnosis, the median age at the first PSA test was 55.8 years.

PSA values ranged from near zero to 22,510 ng/mL, with a median value of 1.2 ng/mL (IQR, 0.7–2.6 ng/mL). Overall, 82% of men who underwent repeated PSA testing never reached the age-specific referral thresholds used in Sweden (3 ng/mL for men younger than 70 years, 5 ng/mL for those aged 70–80 years, and 7 ng/mL for those older than 80 years).

During follow-up, 30,199 men (6.0%) were diagnosed with prostate cancer. A summary of PSA retesting patterns and outcomes by age can be found in Table 1. The median age at diagnosis was 69.3 years (IQR, 63.1–75.7). The median time from the first PSA test to diagnosis was 4.5 years (IQR, 0.0–9.5).

**Table 1.** Summary of PSA testing, biopsy, and prostate cancer diagnosis by age group and calendar period, Stockholm 2003-2021.

| Period | Age group (years) | Person-years (PY) (x1,000) | Tests total (x1,000) | Tests per PY | PSA, Median (IQR), ng/mL | Biopsies total | Biopsies per PY | Diagnosed total | Diagnosed per PY |
| --- | --- | --- | --- | --- | --- | --- | --- | --- | --- |
| 2003–2008 | <50 | 268.39 | 96.96 | 0.36 | 0.70 (0.50-1.00) | 3,050 | 0.011 | 2,192 | 0.008 |
|  | 50–59 | 269.34 | 174.01 | 0.65 | 0.90 (0.60-1.60) | 18,398 | 0.068 | 14,974 | 0.056 |
|  | 60–69 | 265.72 | 190.76 | 0.72 | 1.40 (0.80-2.80) | 32,110 | 0.121 | 23,704 | 0.089 |
|  | 70–79 | 152.73 | 113.46 | 0.74 | 2.50 (1.20-5.00) | 20,335 | 0.133 | 13,907 | 0.091 |
|  | 80+ | 88.39 | 61.51 | 0.70 | 3.40 (1.40-7.90) | 6,838 | 0.077 | 5,120 | 0.058 |
| 2009–2014 | <50 | 585.01 | 121.05 | 0.21 | 0.73 (0.50-1.10) | 2,725 | 0.005 | 1,324 | 0.002 |
|  | 50–59 | 733.51 | 215.78 | 0.29 | 0.97 (0.60-1.60) | 19,278 | 0.026 | 10,661 | 0.015 |
|  | 60–69 | 625.08 | 259.24 | 0.41 | 1.50 (0.81-2.80) | 48,109 | 0.077 | 24,135 | 0.039 |
|  | 70–79 | 274.34 | 140.31 | 0.51 | 2.20 (1.10-4.30) | 29,355 | 0.107 | 13,801 | 0.050 |
|  | 80+ | 133.12 | 66.33 | 0.50 | 3.30 (1.43-7.20) | 9,980 | 0.075 | 4,990 | 0.037 |
| 2015–2021 | <50 | 840.84 | 157.57 | 0.19 | 0.69 (0.47-1.00) | 1,727 | 0.002 | 695 | 0.001 |
|  | 50–59 | 799.40 | 234.64 | 0.29 | 0.94 (0.58-1.60) | 14,053 | 0.018 | 4,965 | 0.006 |
|  | 60–69 | 553.09 | 241.05 | 0.44 | 1.50 (0.80-2.80) | 34,953 | 0.063 | 11,600 | 0.021 |
|  | 70–79 | 405.96 | 198.72 | 0.49 | 2.00 (1.00-3.70) | 37,818 | 0.093 | 11,901 | 0.029 |
|  | 80+ | 154.61 | 72.28 | 0.47 | 2.70 (1.20-6.00) | 12,600 | 0.081 | 3,853 | 0.025 |

### Statistical methods

In outline, we fitted a joint model that linked three processes: the longitudinal trajectory of log-transformed PSA over age, time to prostate cancer diagnosis, and time to PSA test. The longitudinal submodel used a linear mixed-effects model to describe individual log-transformed PSA values as a nonlinear function of attained age, represented using a natural cubic spline with three degrees of freedom. Patient-level random intercepts and slopes were included to allow individual log-transformed PSA trajectories to deviate from the population-average trend.

Time to prostate cancer diagnosis was modelled using a left-truncated proportional hazards model, with the hazard proportional to: (i) a baseline hazard specified as a function of attained age using a natural cubic spline with three degrees of freedom; and to (ii) the underlying log-transformed PSA trajectory defined in the longitudinal submodel.

Time to PSA testing was modelled using a recurrent-event proportional hazards model in a counting-process formulation, with the hazard proportional to: (i) a baseline hazard specified as a function of attained age using a natural cubic spline with three degrees of freedom; and to (ii) the underlying log-transformed PSA trajectory from the longitudinal submodel. This formulation accounts for the correlation among repeated tests within individuals.

Correlations between the three processes were thus modelled through the shared underlying longitudinal PSA trajectory, which was modelled explicitly as a function of attained age. Each submodel can therefore be viewed implicitly as a model of attained age. Additionally, from the fitted joint model, we were able to estimate patient-specific trajectories for all three processes.

To contrast the joint model with modelling each process in isolation, we fitted corresponding standalone models to each component. Longitudinal PSA values were modelled using a linear mixed-effects model. Time to prostate cancer diagnosis was modelled using a left-truncated proportional hazards model. The PSA-testing process was modelled using a recurrent-event proportional hazards model in start–stop form, including a subject-specific frailty term.

A detailed formulation of the joint model is provided in the Supporting information. All analyses were performed using R version 4.4.3, with the joint model fitted using the VAJointSurv package available on CRAN.

## Results

Table 1 provides an overview of the PSA testing patterns. Total PSA values together with rates for PSA testing, biopsies and diagnosis are higher at increasing ages.

### Joint model

Fig 1 displays the population-level estimates with age as the time scale. Looking at the three panels, we can see the three processes that form the core of the joint model: the longitudinal evolution of PSA (left), the age-specific hazard of undergoing a PSA test (middle), and the age-specific hazard of prostate cancer diagnosis (right). The green curves show population-level predictions with 95% pointwise prediction intervals that account for both fixed-effect uncertainty and between-individual variability.

**Fig 1.**
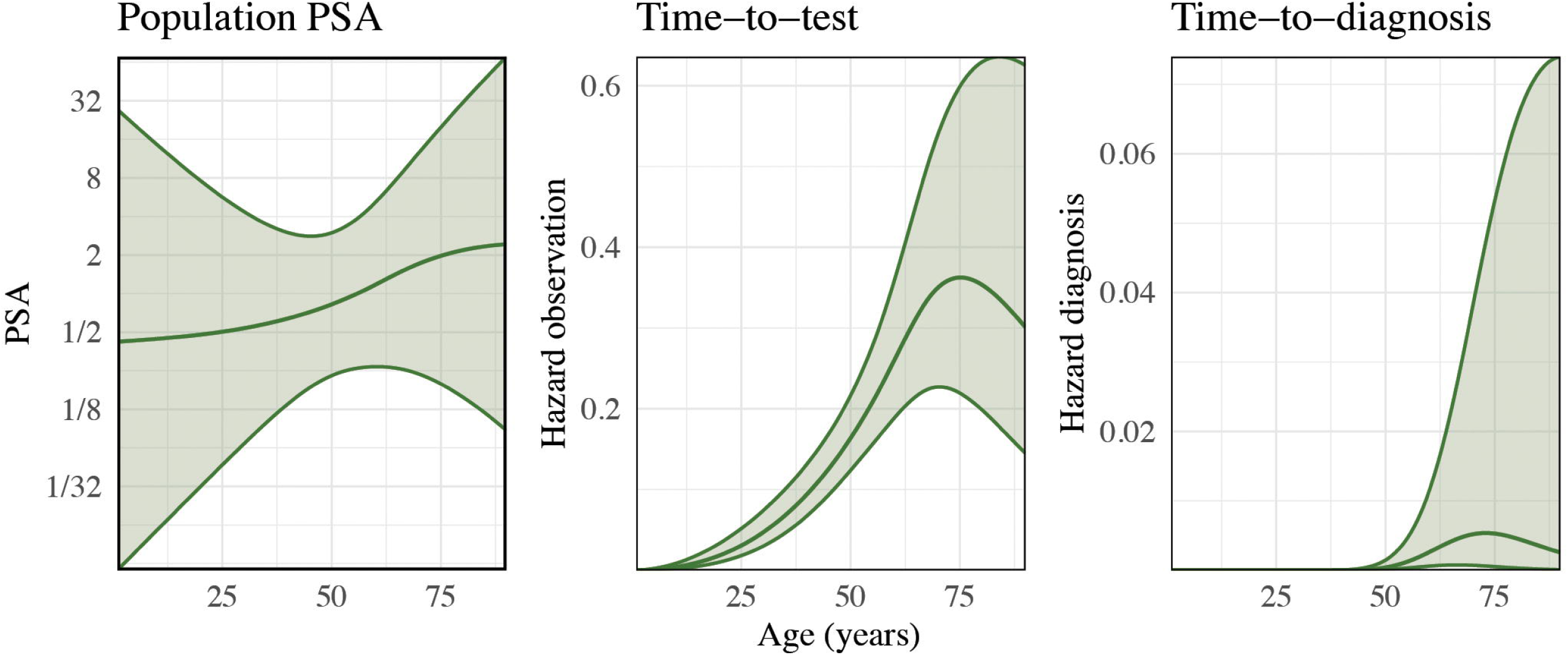
Estimated population-level longitudinal and time-to-event processes from the joint model.

### Longitudinal submodel

The longitudinal submodel showed that population-average log PSA values increased nonlinearly with age. There was substantial heterogeneity in PSA trajectories across men, reflected by the large random-effect variances. The standard deviation of the random slope was 677.95 (95% CI: 674.74–681.17; P < .001), and the standard deviation of the random intercept was 0.812 (95% CI: 0.810–0.815; P < .001). The correlation between the intercept and slope was high at 0.59 (95% CI: 0.59–0.60; P < .001), indicating that men with higher baseline PSA tended to have steeper age-related increases. Additionally, we observed a steady increase in the standard deviation of the residuals (Fig 2), indicating heteroscedasticity. In other words, the magnitude of unexplained variation in log PSA grows larger with age.

**Fig 2.**
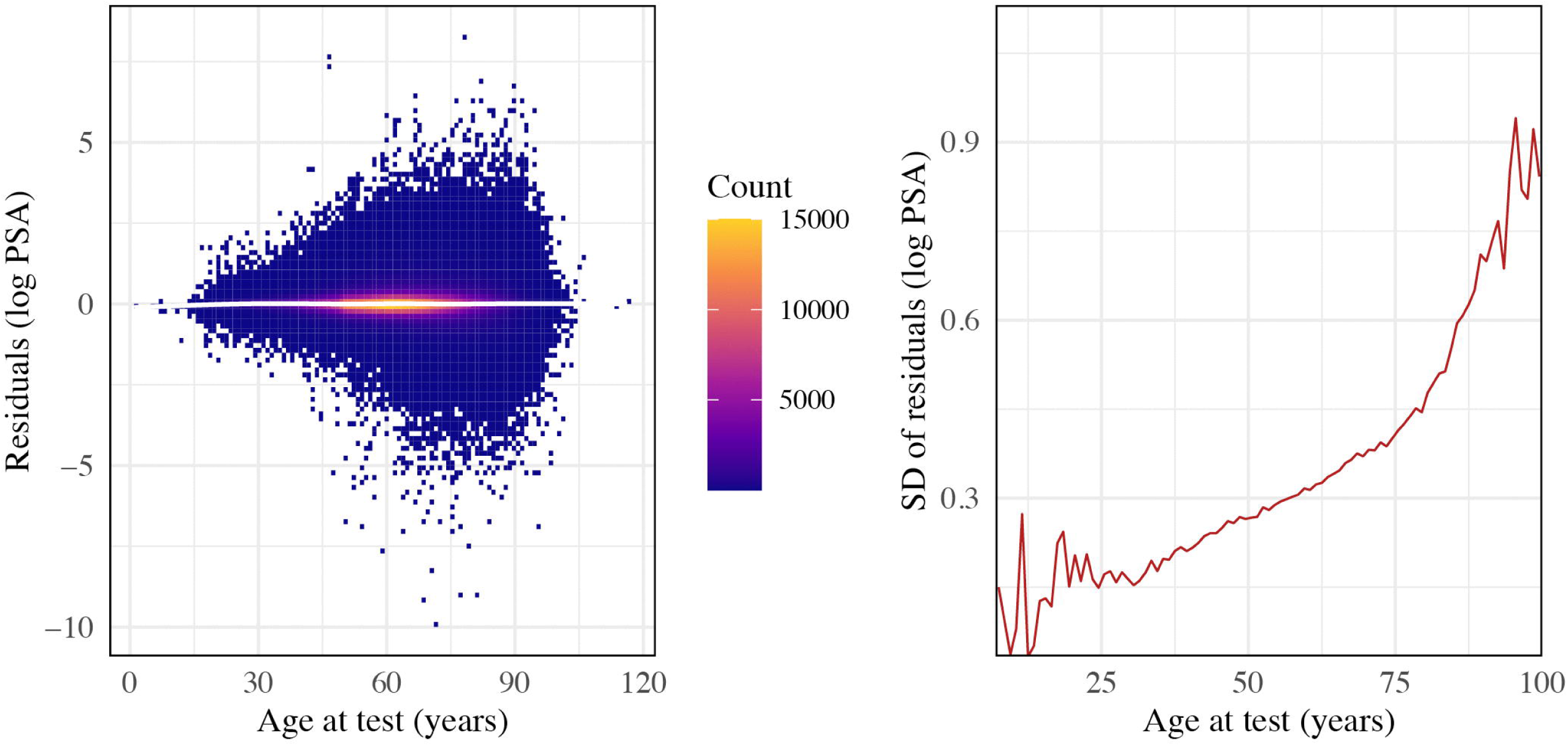
**Heatmap (left) and standard deviations (right) of residuals from the longitudinal component of the joint model for log-transformed PSA by age at test**.

These findings were consistent with the standalone longitudinal model (Fig S4), although the joint model estimated a slightly lower population-average trajectory.

### Time-to-event submodels

A doubling of PSA concentration was associated with a higher hazard of diagnosis, corresponding to a hazard ratio of 2.01 (95% CI: 1.99–2.02; P < .001). This estimated effect was stronger than that obtained when modelling time to diagnosis separately; the Cox proportional hazards model specified using counting-processes formulation with log PSA as a time-updated covariate and robust standard errors clustered by individual, yielded a hazard ratio of 1.61 (95% CI: 1.59–1.62; P < .001) per doubling of PSA.

For the observational process, the joint model indicated that doubling of PSA was positively associated with the hazard of a subsequent test (HR = 1.163; 95% CI: 1.161–1.165; P < .001). This was consistent with estimates from the Prentice-Williams-Peterson (PWP) model fitted in isolation, where a doubling of PSA was associated with a hazard ratio of 1.068 (95% CI: 1.066–1.070; P < .001).

### Predicted trajectories

Figs 3 and 4 illustrate fitted trajectories from the joint model for two individuals who were not diagnosed and diagnosed with prostate cancer, respectively. The green curves are population-level estimates from Fig 1. The orange curves represent individual-specific predictions based on the conditional modes of the random effects, with orange shaded areas representing 95% confidence intervals. For the diagnosed individual (Fig 4), the steadily rising PSA trajectory is accompanied by a sharp increase in both the hazard of subsequent testing and the hazard of diagnosis. In contrast, the undiagnosed individual (Fig 3) had PSA values that closely follow the population-average trajectory, with corresponding hazards for testing and diagnosis remaining consistent with population-average values. The individual-specific confidence intervals narrow across all three processes as more PSA measurements accumulate, most noticeably in the intervals where the tests are taken. It is important to note that these (orange) bands are conditional on the entire set of observed PSA values for that individual. Consequently, uncertainty in the earlier parts of each trajectory (before the first measurement) is informed by future observations, providing a retrospective estimate of the patient’s past.

**Fig 3.**
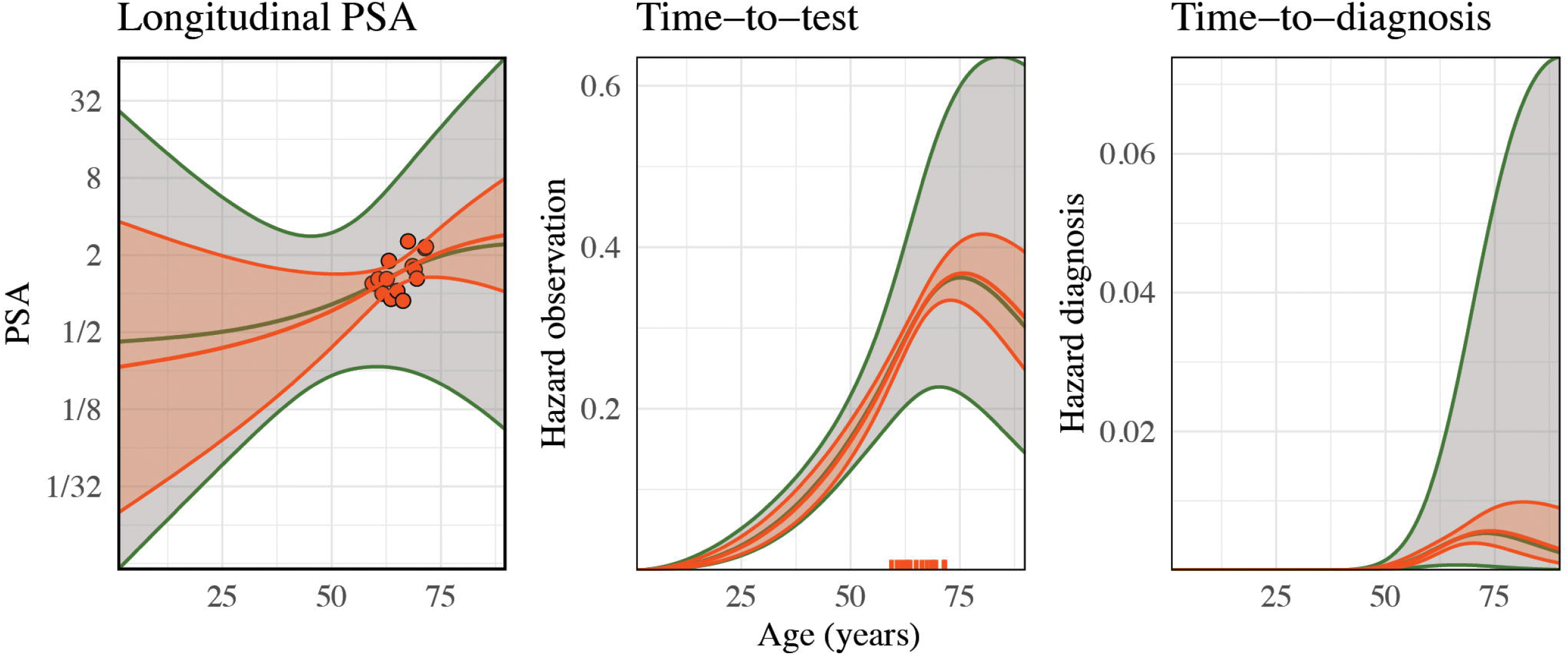
**Estimated longitudinal and time-to-event processes from the joint model for an undiagnosed individual.**

**Fig 4.**
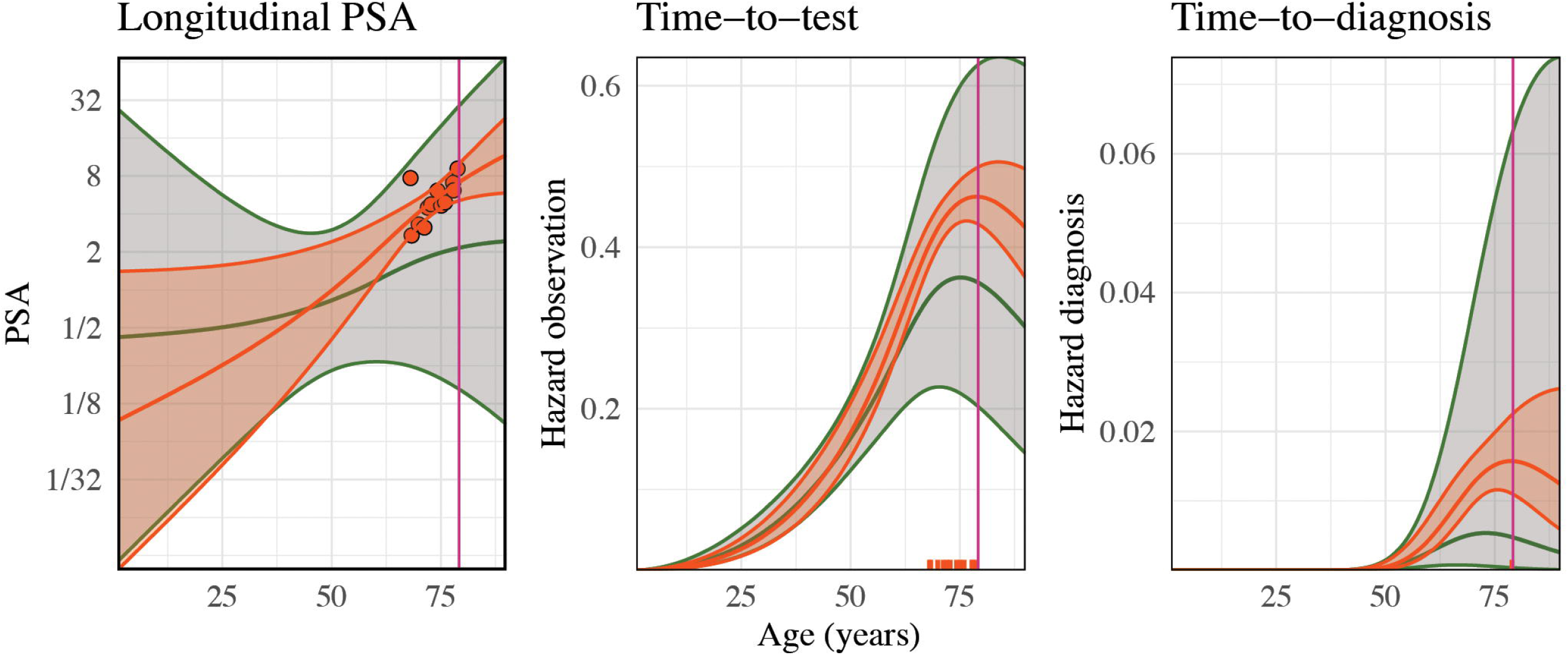
**Estimated longitudinal and time-to-event processes from the joint model for a diagnosed individual; the vertical line represents the age at diagnosis.**

## Discussion

In this large, population-based study of more than half a million men in Stockholm County, we described how PSA values change with age, the observational process of retesting, and the time to prostate cancer diagnosis. Importantly, comparisons with isolated models demonstrated that standard approaches substantially underestimated the association between PSA and diagnosis by failing to account for the interdependence between biomarker trajectory, testing frequency, and cancer risk.

PSA values increased non-linearly with age and were highly heterogeneous across and within men. We observed considerable heteroscedasticity, with the variability in PSA values increasing with age. This pattern partially explains why using a single PSA threshold for screening in large populations is challenging.

Our comparison of the joint model and the linear mixed model for PSA trajectories highlights the importance of accounting for the observation process when analysing clinical biomarkers obtained through opportunistic or patient-driven testing. Men with elevated PSA levels are more likely to be re-tested, leading to an informative observation process in which the timing and frequency of PSA measurements depend on the underlying PSA trajectory. A mixed effects model assumes that measurement times are independent of the outcome, and consequently can lead to biased estimates. By jointly modelling the PSA process and the observation process, the joint model explicitly accounts for this dependency, therefore potentially providing more realistic estimates of population-level PSA trajectories. In our analysis, this adjustment resulted in slightly lower estimated mean PSA levels compared with the linear mixed-effects model, consistent with the expectation that failure to adjust for informative observation tends to overestimate PSA at the population level.

The mean age at diagnosis was 70.8 years, which explains the higher PSA densities observed among men aged 60 and older, likely reflecting cancers that had already developed by that age. However, clear differences in PSA distributions between diagnosed and undiagnosed men were also evident at younger ages.

When the longitudinal PSA process and the event processes were modelled separately, the estimated hazards for diagnosis and retesting were attenuated. For example, the hazard ratio for prostate cancer diagnosis per doubling of PSA was 1.61 in the separate Cox model but increased to 2.01 in the joint model. Similarly, the association between PSA and the risk of retesting rose from HR = 1.068 in the separate PWP model to HR = 1.163 under joint modelling. This attenuation in the separate analyses likely reflects informative observation bias: the timing of PSA measurements is not independent of the biomarker trajectory or the disease progression. When such feedback is ignored, part of the association between PSA and the outcome is absorbed into the unmodelled observation mechanism, leading to an underestimation of effect sizes. The joint model corrects for this by sharing random effects across the longitudinal and survival submodels, therefore accounting for the correlation between PSA levels, testing frequency, and diagnosis risk.

By generating individual-specific predictions with corresponding confidence bounds, the joint model also enables direct assessment of how a man’s PSA trajectory deviates from the population mean, which could help guide subsequent diagnostic steps, such as the need for MRI referral. These conditional predictions quantify both within– and between-person uncertainty, illustrating whether an individual’s observed PSA pattern is consistent with population expectations. Such personalised predictions can be extended to estimate dynamic risks of future testing or diagnosis, integrating an individual’s entire PSA history and testing pattern. In this way, the model provides a framework not only for understanding population-level processes but also for supporting personalised decision-making.

We used variational approximations to fit the joint model. This approach scaled well to a moderately large dataset and supported personalised predictions, including measures of uncertainty.

This study has several limitations. First, we do not explicitly model for a change of PSA trajectory after prostate cancer onset. Although cancer onset is not observable, inspection of individual PSA trajectories suggested that men who were subsequently diagnosed often experienced a more rapid rise in PSA after a certain point. To partially address this issue, we have censored from a prostate cancer diagnosis. Second, the data are observational, leading to substantial variation in the observation process across individuals. Our joint modelling approach seeks to explicitly model for this variation. Third, we lack information on other factors that can elevate PSA, such as urinary tract infections or lower urinary tract symptoms; the absence of these data may contribute to unexplained variability in PSA trajectories. Fourth, our findings may be specific to the Stockholm population. We posit that the connections between the three processes will be similar across populations. It would be useful to perform similar modelling in another population to test this hypothesis.

In future work, we anticipate that this model can be used to estimate a detailed profile for PSA dynamics for use in a prostate cancer screening model, and estimate PSA uptake and rescreening to describe current opportunistic testing in a population. Additionally, we plan to first estimate the cancer onset distribution using a separate screening model, and then incorporate this estimated distribution into a changepoint model for the longitudinal PSA process.

## Conclusions

PSA trajectories, retesting behaviour, and prostate cancer diagnosis are strongly interdependent, and modelling them separately can substantially underestimate associations with PSA. Joint modelling accounts for informative observation in opportunistic testing and enables individual-specific predictions of PSA evolution, retesting, and diagnostic risk. These findings support integrating PSA history and testing intensity into screening models to improve risk stratification and precision.

## Data Availability

Data cannot be shared publicly because of restrictions under the Swedish Public Access and Secrecy Act (Offentlighets-och sekretesslag 2009:400) regarding sensitive patient data. Data are available from the Stockholm Prostate Cancer Diagnostics Register (STHLM0) Steering Committee (contact via the Department of Medical Epidemiology and Biostatistics, Karolinska Institutet) for researchers who meet the criteria for access to confidential data.

