## Supporting information for "Joint modelling of PSA dynamics and prostate cancer risks: A population-based study"

Please note that the main manuscript does not explicitly reference all supporting information.

### S1. PSA retesting seasonal patterns

PSA testing volumes from 2003 to 2020 are shown in Fig S1 and display a consistent seasonal pattern, with sharp declines during the summer months (July–August) and around the winter holidays (December–January). Testing peaks typically occurred in spring and autumn, reflecting the influence of healthcare system scheduling, holiday periods, and patient availability.

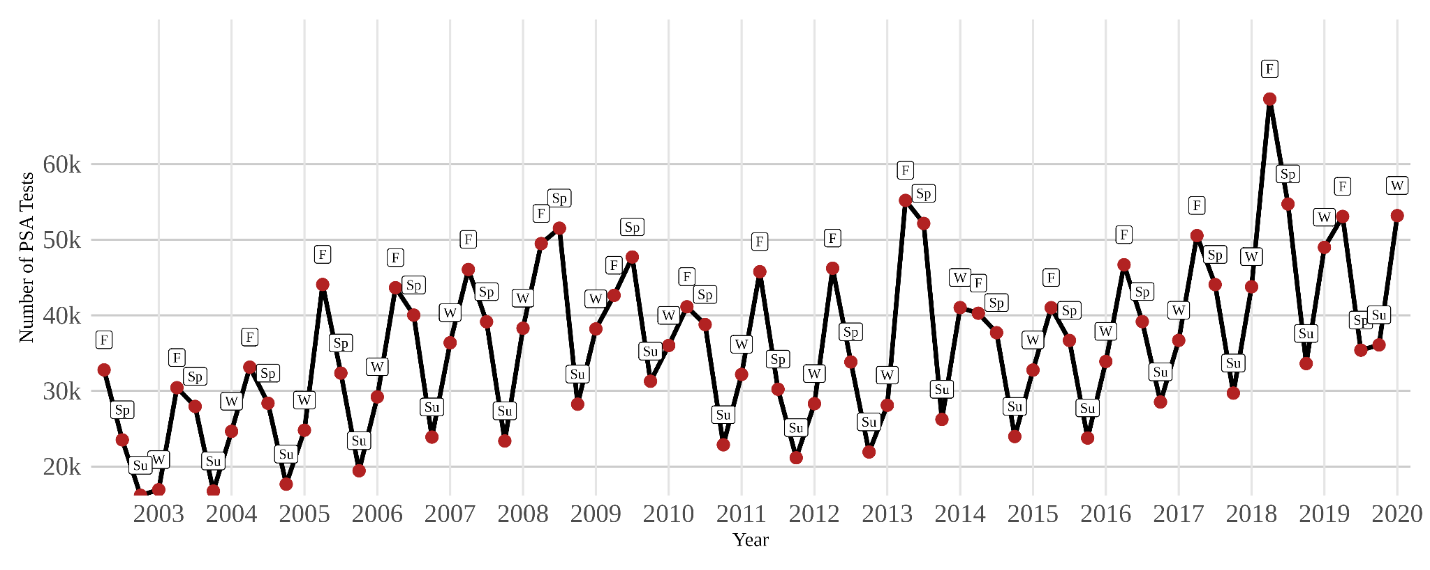

**Fig S1. Number of PSA tests by calendar season and year**

### S2. Longitudinal process for PSA values

PSA values increased with age, and their variance also increased with age; see Fig S2.

The Fig S2 displays that distributions of PSA values had long right tails across all age groups, indicating considerable heterogeneity and the presence of extreme PSA values. In addition, stratifying by diagnosis status showed that log-transformed PSA values were consistently higher among men who were eventually diagnosed with prostate cancer across all age groups (Fig S3).
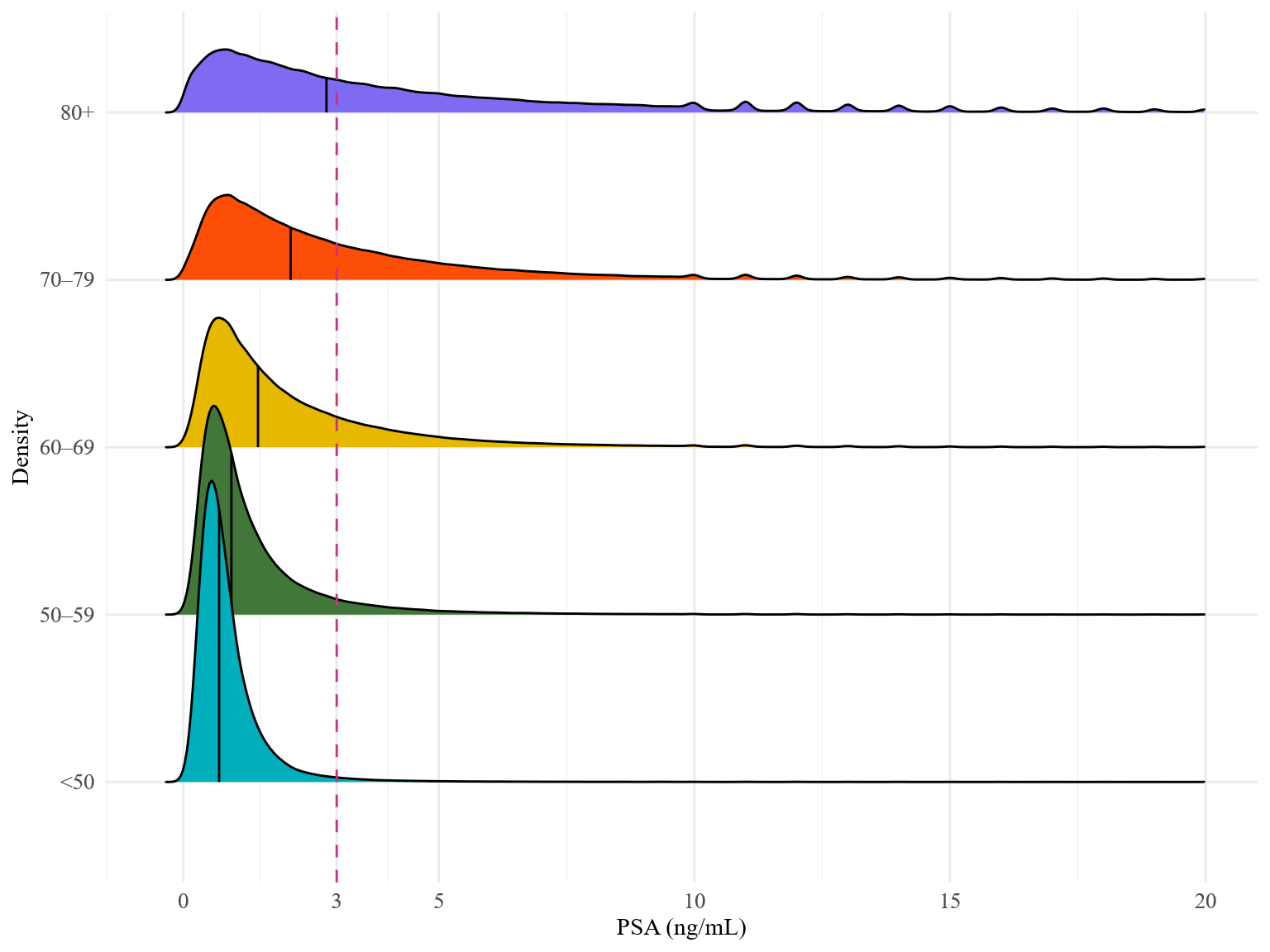

**Fig S2. PSA probability density by age group**, with vertical lines indicating the median values.

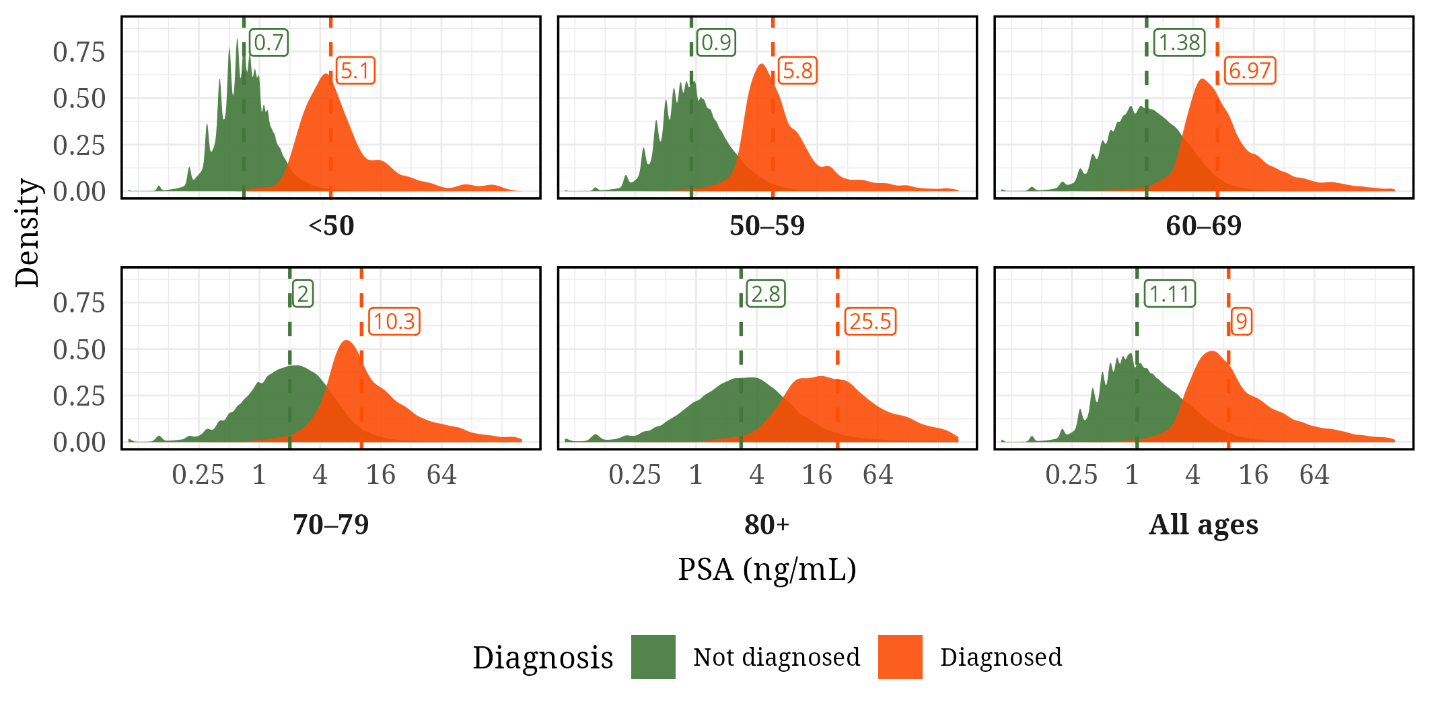
**Fig S3. Distribution of PSA by age group and diagnosis status (x-axis on a logarithmic scale).** For men diagnosed with prostate cancer, PSA measurements were restricted to tests obtained within 90 days prior to diagnosis. Vertical dashed lines indicate group-specific medians; box labels show the corresponding median PSA (ng/mL). Probability densities were estimated on the log PSA scale, but axis labels are displayed on the PSA scale for interpretability.

In a linear mixed-effects model of log-transformed PSA with age modelled by a natural cubic spline with three degrees of freedom, log PSA increased nonlinearly with age (all spline terms, <0.001). The fixed effects log PSA population mean is presented in Fig S4. The residual standard deviation was 0.42, showing considerable within-person variability.

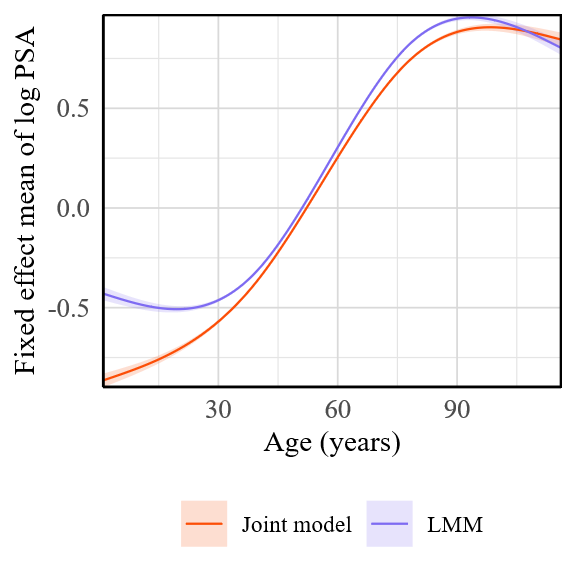

**Fig S4. Comparison of fixed-effect PSA trajectories from the joint model and the linear mixed model.**

### S3. Observation process

In the PWP gap-time model, doubling of PSA was associated with an increased hazard of subsequent testing (HR = 1.12; 95% CI = 1.12 to 1.12; p < 0.001).

Fig S5 represent the cumulative conditional probability of undergoing the next PSA test within 4 years since the previous test, given the individual has taken all preceding tests.

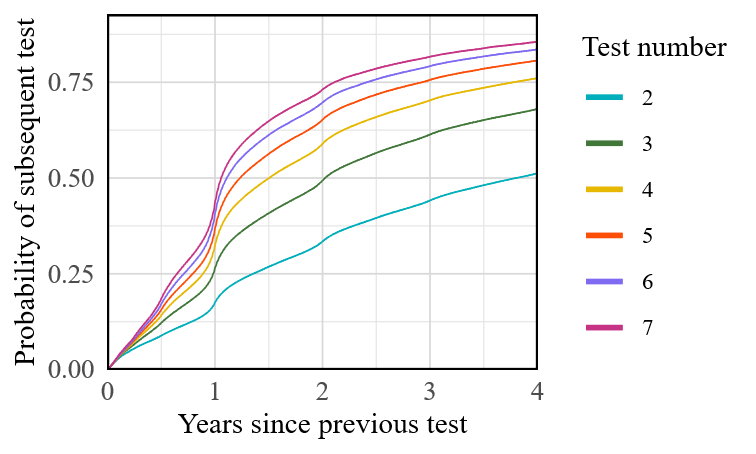

**Fig S5. Cumulative conditional probability of subsequent PSA testing by time since the previous test, stratified by test number.**

### S4. Disease progression

Left-truncated Kaplan–Meier curve for time to prostate cancer diagnosis, conditional on having at least one PSA test in the Stockholm region during 2002-2020 could be found in the Fig S6.

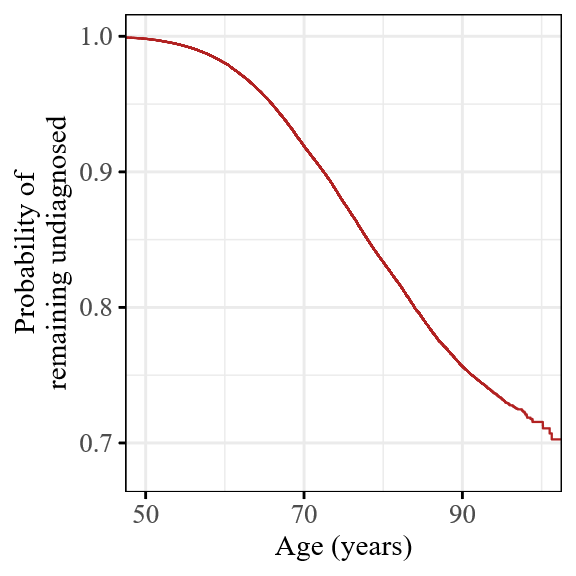

**Fig S6. Kaplan–Meier estimates of the probability of remaining undiagnosed.**

In the Cox proportional hazards model without time splitting we observed evidence of non-proportional hazards (Fig S7). The piecewise Cox model, which allowed the effect of log PSA to vary across age bands, is summarised in Fig S8. The full table of hazard ratios with 95% confidence intervals is provided in Table S1.

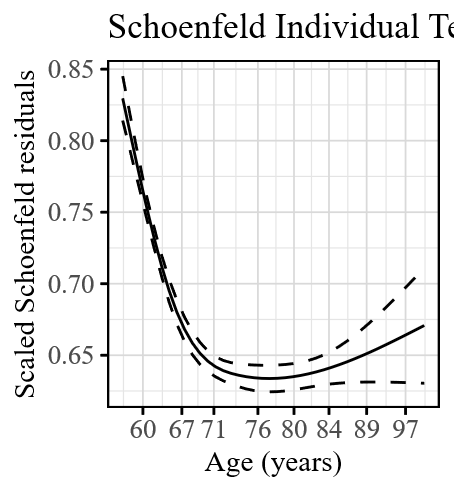

**Fig S7. Schoenfeld residuals and time-varying coefficient for log PSA from the Cox model for time to prostate cancer diagnosis.**

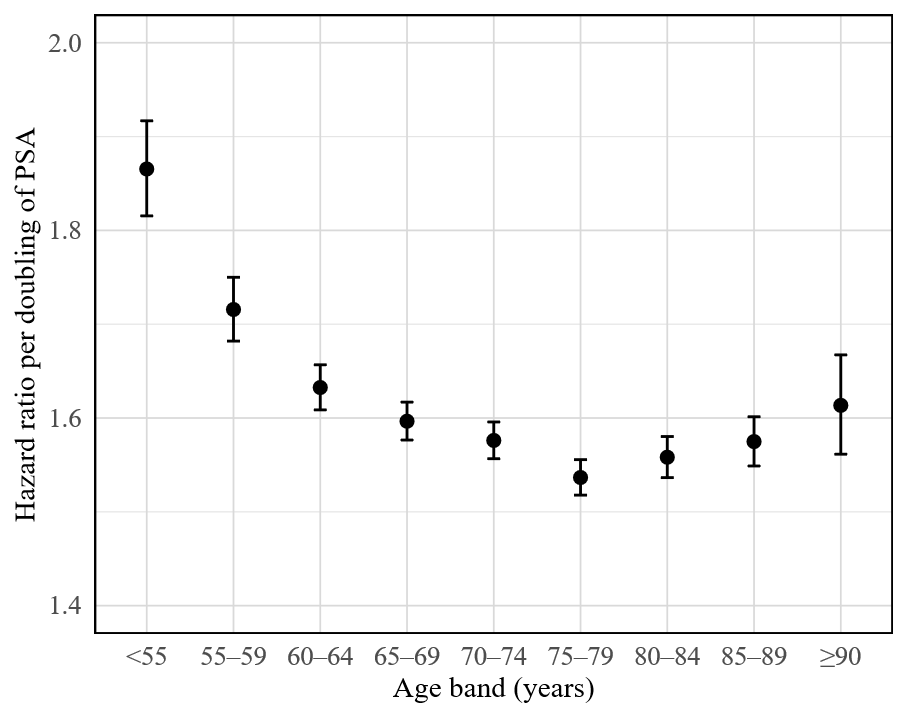

**Fig S8. Age-specific hazard ratios per doubling of PSA from a piecewise Cox model for time to prostate cancer diagnosis.**

| Age group | HR (95% CI) | P-value |
| --- | --- | --- |
| <55 | 1.865 (1.815-1.917) | <0.001 |
| 55–59 | 1.716 (1.682-1.750) | <0.001 |
| 60–64 | 1.633 (1.609-1.657) | <0.001 |
| 65–69 | 1.597 (1.577-1.617) | <0.001 |
| 70–74 | 1.576 (1.557-1.596) | <0.001 |
| 75–79 | 1.537 (1.518-1.556) | <0.001 |
| 80–84 | 1.558 (1.536-1.580) | <0.001 |
| 85–89 | 1.575 (1.549-1.601) | <0.001 |
| ≥90 | 1.613 (1.561-1.667) | <0.001 |

**Table S1. Age-specific hazard ratios per doubling of PSA from a piecewise Cox model for time to prostate cancer diagnosis.**

### Joint model formulation

#### Longitudinal submodel

Let individual $i\in\left[ 1,n \right]$ have $j\in\left[ 1,m_{i} \right]$ PSA tests during the follow-up. Denote the $\text{log}$ PSA value for individual $i$ at the $j$ th test, taken at age $t_{ij}$, by $Y\left( t_{ij} \right)$. The longitudinal submodel is

$$\boldsymbol{Y}\left( t_{ij} \right)=\boldsymbol{X}_{m}\left( t_{ij} \right)\boldsymbol{\beta}_{m}+\boldsymbol{Z}_{m}\left( t_{ij} \right)\boldsymbol{b}_{m}+\epsilon_{ij},$$

where $\boldsymbol{X}_{m}$ is the fixed-effects design matrix, with $\boldsymbol{X}_{m}\left( t_{ij} \right)$ representing the row corresponding to individual $i$ at his $j$th test, $\boldsymbol{\beta}_{m}$ is the vector of fixed-effects parameters, $\boldsymbol{Z}_{m}$ is the random-effects design matrix, and $\boldsymbol{b}_{i}$ are the individual-specific random effects. We assume $\boldsymbol{b}_{i}\sim N\left( \boldsymbol{\mu},\boldsymbol{\Sigma} \right)$ with mean vector $\boldsymbol{\mu}$ and covariance matrix $\boldsymbol{\Sigma}$, and $\epsilon_{ij}\sim N\left( 0,\sigma^{2} \right)$. Disregarding the sharing of random effects with the survival components, this is a standard linear mixed-effects model.

In our application, we specify $\boldsymbol{X}_{m}$ to include an intercept and a natural cubic spline basis for age at test, with two internal knots placed at the tertiles of the observed age-at-test distribution. The random‐effects included a random intercept and a random slope for age, with $\boldsymbol{b}_{i}$ following two-dimensional normal distribution.

#### Time-to-event submodels

The hazard functions at age $t_{ij}$ are defined as

$$h_{k}\left( t_{ij} \right)=\text{exp}\left( \boldsymbol{X}_{s_{k}}\left( t_{ij} \right)\boldsymbol{\beta}_{s_{k}}+\alpha_{k}\boldsymbol{Z}_{m}\left( t_{ij} \right)\boldsymbol{b}_{m} \right),$$

where $k=1$ corresponds to the hazard of diagnosis and $k=2$ corresponds to the intensity of the next PSA test. $\boldsymbol{X}_{s_{k}}$ is the design matrix for the fixed effects, $\boldsymbol{\beta}_{s_{k}}$ is the vector of fixed‐effects parameters, and $\alpha_{k}$ represents the association parameter linking the longitudinal random effects to each hazard process. The models account for left-truncation to adjust for the delayed entry of individuals at varying ages. Disregarding the sharing of random effects through $\alpha_{k}$, these are standard proportional hazards models with normal frailties.

In our application, $\boldsymbol{X}_{s_{k}}\left( t \right)$ is specified similarly to the longitudinal submodel: it includes an intercept and a natural cubic spline basis for age at test, with two internal knots located at the tertiles of the distribution of observed event and censoring times.

The model was fitted using a frequentist full maximum likelihood approach. To approximate the intractable integrals over the random effects, we used Gaussian variational approximations [1]. Technical details of our novel implementation are provided in a separate paper [2].
